# Combining polygenic risk scores to understand genetic liability to physical-mental health multimorbidity in UK BioBank

**DOI:** 10.1101/2025.10.20.25338357

**Authors:** Daniel Stow, Ruby S. M. Tsang, Ioanna Katzourou, Jack F. G. Underwood, The LIfespaN multimorbidity research Collaborative (LINC), Peter Holmans, Inês Barroso, Hilary Martin, Marianne B.M. van den Bree, Sarah Finer, Nic Timpson

## Abstract

**Background:** Internalising and CardioMetabolic MultiMorbidity (ICM-MM) is a common form of mental-physical health multimorbidity, yet its genetic predisposition is largely unknown. We examined the polygenic nature of ICM-MM by assessing single trait-specific polygenic risk scores (PRS_TRAIT_) and whether combining them could increase the proportion of variance in liability to ICM-MM explained by genetic variation.

**Methods:** We developed PRS_TRAIT_ using PRS-CS and summary statistics from the largest trait-specific GWAS excluding UK Biobank (UKB). We evaluated PRS_TRAIT_ on ICM-MM risk in 206,452 UKB participants (n=39,311 (19.0%) with ICM-MM) using logistic regression adjusted for gender and 10 genetic principal components, defining ICM-MM as lifetime occurrence of: ≥1 internalising (depression, anxiety, somatoform disorder) traits AND ≥1 cardiometabolic traits (type 2 diabetes, obesity, hypertension, dyslipidemia, chronic kidney disease). We trained an elastic net in a 50% subsample to generate ICM-MM-PRS_TRAIT_: a weighted combination of PRS_TRAIT_ targeting ICM-MM.

**Results:** The strongest associations were between ICM-MM and PRS_TRAIT_ for depression and type 2 diabetes - both odds ratios (OR) 1.18, [95% confidence interval (CI) 1.17–1.20] per standard deviation increase in PRS_TRAIT_. ICM-MM-PRS_TRAIT_ retained five PRS_TRAIT_ with stronger associations (OR=1.31, [95%CI 1.29–1.34]) than any PRS_TRAIT_ in the validation sample.

**Discussion:** Combining several PRS explains more variance in ICM-MM liability than single-trait PRSs alone. ICM-MM-PRS_TRAIT_ is a measure of genetic risk that could be used to examine premorbid stages of ICM-MM in external and youth cohorts, supporting awareness of earlier presentation and potentially avoidance or intervention.

## Introduction

Multimorbidity, the co-occurrence(1,2) or lifetime occurrence(3) of two or more long-term health conditions, is a major public health concern, affecting over a third of the global population in mid-life(4,5). Multimorbidity prevalence is rising,(6,7) leading to increased personal,(8,9) societal, and healthcare costs.(10,11)

The co-occurrence of internalising (depression, anxiety) and cardiometabolic conditions (hypertension, obesity, type 2 diabetes) is a common form of mental and physical healthy multimorbidity.(12–15) The interaction between mental health conditions and physical multimorbidity increases unplanned (16) and emergency department (17) hospital admissions, and accounts for a substantive proportion of total secondary care costs in England.(18)

Evidence for intervention and management of multimorbidity is growing (19,20) but little is known about prevention or early life manifestations of multimorbidity.(21) Preventing or delaying onset of multimorbidity would have major benefits for affected individuals, as well as for health care provision and costs.(22) Genetic variation has been implicated in the development of internalising and cardiometabolic disorders, including rare and common factors.(23–31) These common, complex disorders have a polygenic architecture and an individual’s susceptibility to disease can partly be captured by polygenic risk scores. Polygenic risk scores can be used to identify subgroups with increased risk of disease and predict case status in both case-control and population-based cohort studies.(32)

Recent studies have shown that clinical manifestations of polygenic risk emerge early in life, for example, significant gradients of polygenic risk emerged in early childhood for obesity and cardiometabolic traits.(33,34) Understanding how genetic risk of ICM-MM manifests in early life could support the development of tailored early intervention and prevention strategies to delay or prevent multimorbidity in later life, but the evidence on polygenic scores to support this undertaking are lacking.

### AIMS

The aims of this study were to identify a polygenic risk score for ICM-MM by i) evaluating associations between single trait-specific PRSs and lifetime ICM-MM risk, and ii) examining whether a combination of existing scores could increase the proportion of variance in liability to lifetime ICM-MM explained by genetic variation.

## Materials and Methods

This work was conducted as part of the Lifespan Multimorbidity Research Collaborative (LINC)(35), which studies development of internalising and cardiometabolic multimorbidity (ICM-MM) across the lifespan in five longitudinal population-based cohorts with genetic data.

### Patient and public involvement

Members of the public with lived experiences of multiple long-term conditions have been involved throughout the course of the LINC study. For this study they assisted primary and secondary care doctors, and other members of the research team to select the conditions that make up the ICM-MM phenotype (see below “phenotype definition”) and informed the research questions and aims.

### Ethics

UK Biobank has approval from the North West Multi-centre Research Ethics Committee (reference 16/NW/0274) as a Research Tissue Bank. Participants provided electronic signed consent at recruitment.

### Data source

UK Biobank (UKB) is a cohort comprising over 500,000 people living in the United Kingdom. Participants aged 40 to 69 years old were recruited between 2006 and 2010. Participants attended a baseline assessment and were also invited to attend repeat assessments over the following ten years. Our study was conducted under application number 79704.

### Participants

Participant recruitment processes and eligibility criteria for UKB are described in detail in Bycroft et al., 2018.(36) We restricted our analysis to the subset of individuals with primary care electronic healthcare record (EHR) data (∼230,000, or 45% of the UKB cohort)(37).

### Phenotype definition

LINC investigates a specific form of multimorbidity: internalizing and cardiometabolic multimorbidity (ICM-MM), defined as the lifetime occurrence of at least one internalising condition (anxiety, depression, somatoform disorder) AND at least one cardiometabolic condition (chronic kidney disease, dyslipidemia, hypertension, obesity, type 2 diabetes). The cardiometabolic conditions were selected because they occur relatively early on the pathways to cardiovascular disease with divergence in risk starting early in life,(38–40) rendering them suitable for study across the lifespan.

In this study, we used UKB electronic health record data (primary care and NHS Digital Hospital Episode Statistics) to identify participants with ICM-MM using existing SNOMED and ICD-10 codelist resources(41)(42). The presence of any one (or more) code(s) was taken as a diagnosis of the condition.

### PRS construction

**FIGURE 1**

**FIGURE 1:**
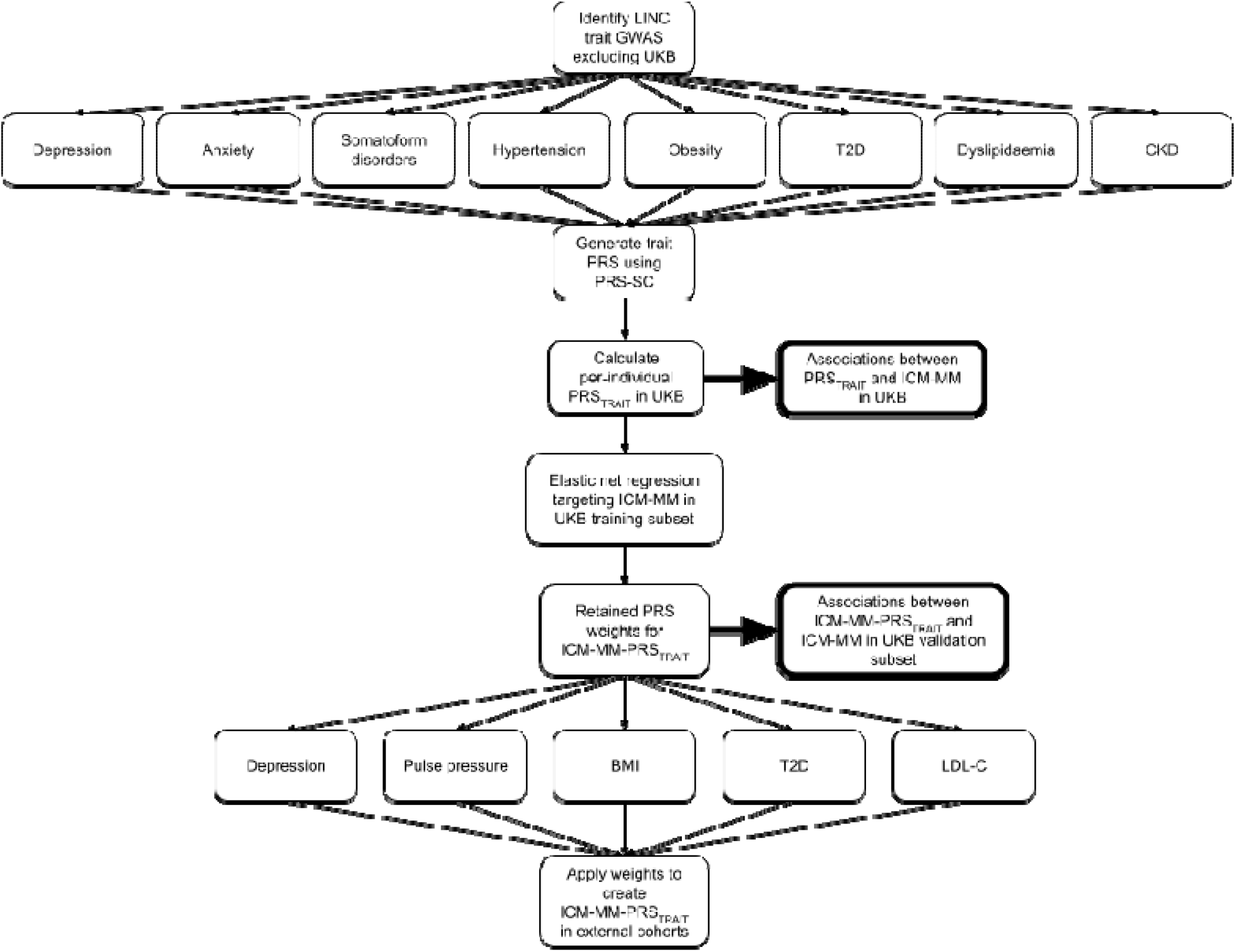
Methods flowchart for generating and testing the PRSs. GWAS(genome wide association study), BMI (body mass index, a proxy for obesity), T2D (type 2 diabetes), LCL-C (LDL-cholesterol, a proxy for dyslipidaemia), CKD (chronic kidney disease), PRS (polygenic risk score), ICM-MM (Internalising and CardioMetabolic MultiMorbidity).

### Constructing trait specific PRSs

Figure 1 summarises the methods to identify the polygenic risk scores and test their associations with ICM-MM. We searched existing literature and the PGS catalogue(43,44) to identify PRS for the eight ICM-MM traits: ‘PRS_TRAIT_’. To avoid overfitting in our UKB analysis, we restricted our searches to PRS constructed using non-UKB data.

If we could not find an existing PRS for ICM-MM conditions meeting these criteria, we searched for GWAS summary statistics from non-UKB studies, or meta-GWAS where authors reported or made available summary statistics excluding UKB. If our searches identified multiple eligible PRS or GWAS for an ICM-MM condition, we selected the one with the largest number of participants. Once appropriate GWAS summary statistics had been obtained, we used PRS-CS(45) and the European ancestry 1000-genomes external linkage disequilibrium (LD) reference panel to calculate PRS. PRS-CS uses a Bayesian algorithm to infer posterior SNP effect sizes via continuous shrinkage (CS), avoiding the reduction in effective sample size for downstream analysis that arises from linkage disequilibrium pruning and p-value thresholding methods.(45) We used PLINK 2.0(46,47) to calculate per-individual PRS_TRAIT_ using the inferred posterior SNP effect sizes and the UKB genotype/array data.

### UKB genotype quality control

Details of the standard quality-control performed on UKB’s genotype data are available in Bycroft et al, 2018.(36) We further removed variants with a low INFO score (<0.9), high missingness (>0.05), low minor allele frequency (<0.01) or variants departing from Hardy–Weinberg equilibrium (p < 10−6), and individuals with high missingness (>0.05) or sex discordance using PLINK 2.0.(46,47) Sex chromosomes were excluded. We used Kinship-based INference for GWAS (KING)(48) to identify individuals related to the second-degree(*r*^2^ > 0.0884), and removed one individual from each related pair at random. After quality control, 449,646 individuals and 6,899,626 variants were retained.

### Constructing an ICM-MM specific PRS

We used elastic net regression, implemented in PRSmix(49) to produce a linear, weighted combination of ICM-MM trait-specific PRS_TRAIT_ (ICM-MM-PRS_TRAIT_) that best predicted ICM-MM. PRSmix methods are described in detail in Truong et al., 2024.(49) Briefly, PRSmix employs elastic net regression on a random subset of training data (specified here as 50%) to identify the optimal, weighted linear combination of input PRSs for predicting the presence of the target trait. The input model was additionally adjusted for self-reported gender and 10 genetic principal components. PRSmix produces information on predictive performance (odds ratios [OR] and area under the curve [AUC]) for the new, weighted combination of PRS in the remaining 50% testing set, compared to i) the ‘null’ input model (adjusted for gender, and 10 genetic principal components only), and ii) the best performing *single* PRS. All PRSs were standardised (z-transformation) before running PRSmix.

### Statistical analysis

We used logistic regression to examine the association between each PRS_TRAIT_, lifetime internalising or cardiometabolic traits and ICM-MM. All models were adjusted for self-reported gender and 10 genetic principal components to account for population stratification and ancestry. We also report area under the curve (AUC) from receiver operating characteristic curves for these logistic models to provide estimates of discrimination between cases and controls. We corrected analyses for multiple testing via Bonferroni correction.

### Supplemental analysis

Given the genetic correlation between the conditions used by LINC to define ICM-MM (see above ‘phenotype definition’) and other cardiometabolic conditions such as cardiovascular disease and ischaemic stroke; and to further test the potential to combine existing polygenic risk scores to predict ICM-MM, we carried out a supplemental analysis using polygenic risk scores calculated by Genomics PLC (PRS_GPLC_) and available under category 300 in the UKB data showcase. SNP weights are not publicly available, but the ‘enhanced’ PRS_GPLC_ have been calculated directly in UKB and select additional external cohorts. PRS_GPLC_ have been constructed for a range of traits, including many of the ICM-MM cardiometabolic traits, but not for any of the internalising ICM-MM traits (see Thompson et al (50) for further details).

More detailed methods for how the Genomics PLC PRS were derived may be found in Thompson et al, 2022,(51) and Thompson et al., 2024.(50) Briefly: Genomics PLC used existing GWAS and proprietary methodologies to generate two sets of PRS for a range of binary and quantitative traits in UKB. The ‘standard’ PRS set was generated for 39 binary and quantitative and binary traits for all 500K UKB participants. The ‘standard’ set uses per-trait meta-analysis GWAS from studies *excluding* UKB (i.e. trained externally). An ‘enhanced’ set of PRSs was also generated for a mixed-ancestry UKB subgroup comprising n=∼120k participants, using scores from the original per-trait meta GWAS (used to develop the ‘standard’ scores), meta analysed with GWAS from the ‘white British unrelated’ subset of UKB (n=∼337k) for 51 binary and quantitative traits (i.e. external training data + UKB subset training data). Genomics PLC have calculated PRSs for participants in external cohorts using the weights derived from the ‘enhanced’ set. If the external cohort contributed information to the ‘standard’ score meta-GWAS, Genomics-PLC re-ran their PRS generation pipeline, excluding the cohort from the meta-GWAS.

We calculated the correlations between each individual PRS_TRAIT_ and ‘enhanced’ PRS_GPLC_ to assess how similar these were. We then repeated the logistic regression, ROC, and PRSmix analyses outlined previously using PRS_GPLC_ to create ICM-MM-PRS_GPLC_. We also repeated the PRSMix analysis with PRS_TRAIT_ as inputs using the same training and testing subset used to create ICM-MM-PRS_GPLC_, and compared the AUC for ICM-MM-PRS_TRAIT_ and ICM-MM-PRS_GPLC_ using DeLong’s test.(52) All models were adjusted for gender and 10 genetic principal components.

### Software

All analyses were performed using R Statistical Software(53) on the UKB Research Access Platform. For ROC analyses we used the pROC package(54)

## Results

**FIGURE 2**

**FIGURE 2:**
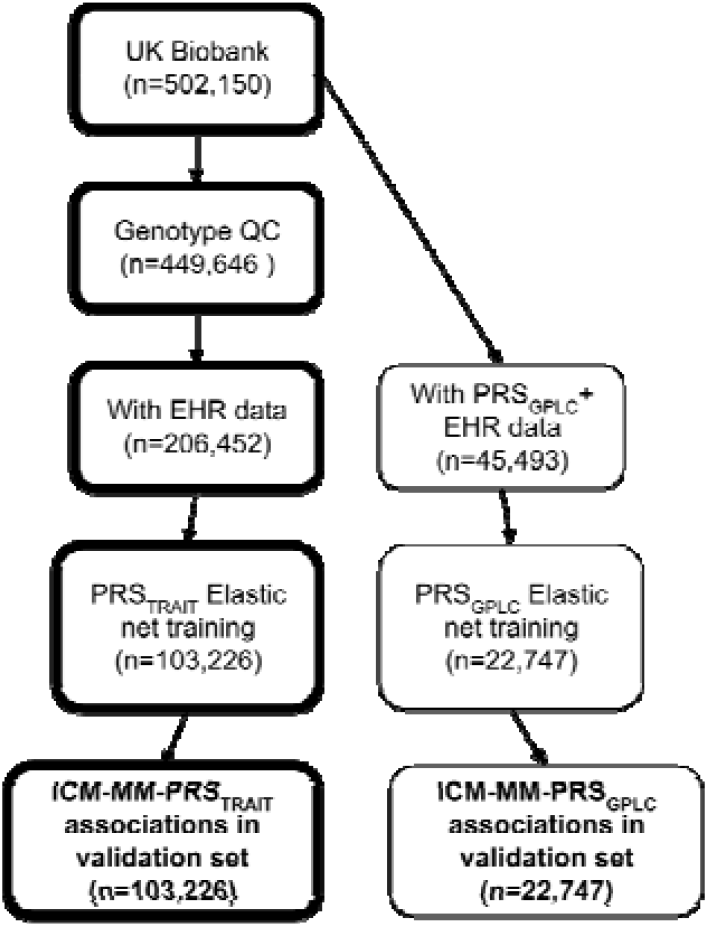
Flowchart of participants at each stage of analysis

### Participants

Figure 2 shows the number of participants at each stage of the analysis. We included data from 206,452 eligible UKB participants (n=111,953 [54.2%] female, mean age of recruitment 66.3 [sd=8.0] years) with electronic healthcare data and genotype data passing QC.

### Identifying trait specific PRSs

Our searches identified no eligible pre-existing PRS for ICM-MM conditions. Characteristics of eligible GWAS for depression,(23) anxiety, (55)T2D,(56) CKD,(57) BMI, (58) LDL-Cholesterol,(59) (used here as a proxy for dyslipidaemia) and diastolic/systolic/pulse pressure(60) (used here as a proxy for hypertension) are in Supplemental table 1. We could not identify any eligible GWAS for somatoform disorders.

### Associations between trait-specific PRS and ICM-MM

Table 1 shows odds ratios (OR) for each PRS_TRAIT_ ICM-MM. The strongest associations with ICM-MM were for PRSs for type 2 diabetes (OR=1.18, 95%CI:1.17 - 1.20) and major depression (OR=1.18, 95%CI:1.17 - 1.20).

**TABLE 1:** The association between LINC ICM-MM trait-specific PRSs and ICM-MM.

| Original PRS<sub>TRAIT</sub> target | AUC-PRS<sub>TRAIT</sub> (95%CI) | Nagelkerke R<sup>2</sup> | OR-PRS<sub>TRAIT</sub> (95%CI) | P value |
| --- | --- | --- | --- | --- |
| Depression | 0.56 (0.56-0.56) | 0.011 | 1.18 (1.17-1.20) | 3.56x10<sup>-153</sup> |
| T2D | 0.56 (0.55-0.56) | 0.010 | 1.18 (1.17-1.20) | 2.18x10<sup>-117</sup> |
| BMI | 0.55 (0.55-0.56) | 0.009 | 1.13 (1.11-1.14) | 4.47x10<sup>-76</sup> |
| Systolic BP | 0.55 (0.55-0.56) | 0.009 | 1.14 (1.12-1.16) | 3.38x10<sup>-73</sup> |
| Diastolic BP | 0.55 (0.55-0.56) | 0.009 | 1.14 (1.13-1.16) | 2.41x10<sup>-75</sup> |
| Pulse pressure | 0.55 (0.55-0.56) | 0.009 | 1.14 (1.13-1.16) | 8.78x10<sup>-75</sup> |
| LDL-C | 0.55 (0.54-0.55) | 0.007 | 1.06 (1.05-1.07) | 7.44x10<sup>-25</sup> |
| Anxiety | 0.55 (0.55-0.55) | 0.007 | 1.11 (1.09-1.13) | 1.05x10<sup>-39</sup> |
| CKD | 0.54 (0.54-0.55) | 0.006 | 1.02 (1.01-1.04) | 0.001 |

Odds ratios (OR) and AUC for each PRS<sub>TRAIT</sub> from logistic regression adjusted for self-reported gender and 10 genetic PCs in 206,452 UKB participants. PRS (polygenic risk score), AUC (area under the curve), CI (confidence interval), OR (odds ratio), T2D (type 2 diabetes), BMI (body mass index), BP (blood pressure), LDL-C (low density lipoprotein-C), CKD (chronic kidney disease)

**TABLE 1**

Supplemental figure 1 has metrics for the internalising and cardiometabolic conditions. Overall, the strongest associations were in the cardiometabolic cluster of conditions (e.g. ORs all >1.3 for PRSs_TRAIT_ for blood pressure measures vs cardiometabolic conditions) and strong associations in the cardiometabolic cluster of conditions attenuated in the ICM-MM models (e.g. association of PRS_TRAIT_ for T2D with cardiometabolic conditions was stronger than for the association PRS_TRAIT_ for T2D with ICM-MM).

### Associations between ICM-MM specific PRS and ICM-MM

In the validation dataset (n=103,226), the OR for ICM-MM-PRS_TRAIT_ was 1.31 (95%CI:1.29-1.34) and the AUC was 0.57 (95%CI 0.57-0.58) - higher than the strongest PRS_TRAIT_ - ICM-MM association, which was for major depression OR=1.19 (95%CI:1.16-1.21), AUC=0.56 (95%CI:0.55-0.56). Details of the five retained PRS_TRAIT_, and optimized weights are shown in Supplemental figure 2. The ICM-MM-PRS_TRAIT_ model had a significantly higher AUC than the model adjusted for major depression (DeLong’s test p-value < 2.2e-16), and both scores improved predicted performance versus the null model (adjusted for gender and 10 genetic PCs only, AUC=0.53, 95%CI=0.53-0.54)

### Supplemental analyses

Genomics PLC ‘enhanced’ polygenic risk scores (PGS_GPLC_) were available for n=45,493 UKB participants with primary care data. Correlations between PGS_TRAIT_ and PGS_GPLC_ are shown in Supplemental figure 3 - The highest correlation for PGS_TRAIT_ and PGS_GPLC_ was for LDL-cholesterol (*r =* 0.55).

PGS_GPLC_ for hypertension (OR=1.25 95%CI:1.22-1.28) and ischaemic stroke (OR= 1.25, 95%CI:1.22-1.28) were most strongly associated with ICM-MM, and had the largest AUC (0.58, 95%CI: 0.57-0.59, and 0.58, 95%CI:0.57-0.58, respectively). For ORs and AUC for all PGS_GPLC_ please see Supplemental figures 4 and 5

Elastic net regression retained 13 of 51 PGS_GPLC_. Details of the retained PRS, and optimized weights for the linear combination are shown in Supplemental figure 6 . OR for ICM-MM-PGS_GPLC_ was 1.32 (95%CI:1.28-1.37) and AUC was 0.59 (95%CI 0.58-0.60). The best performing single PGS_GPLC_ in the validation set (n=22,747) was for ischaemic stroke (OR=1.24, 95%CI: 1.20-1.28, AUC=0.57, 95%CI:0.56-0.58). Both scores showed improved predicted performance versus the null model (adjusted for gender and 10 genetic PCs only, AUC=0.54, 95%CI=0.53-0.55).

In this subset of data, Elastic net regression retained four of seven PRS_TRAIT_ (Supplemental figure 7) and OR for ICM-MM-PRS_TRAIT_ in this subset was 1.27 (95%CI:1.21-1.34) and AUC was 0.56 (95%CI:0.55-0.57). DeLong’s test demonstrated that the models adjusted for ICM-MM-PRS_GPLC_ and PRS_GPLC_ for ischaemic stroke had significantly higher AUC than for ICM-MM-PRS_TRAIT_ (p=8.137e-09 and p=0.027 respectively).

## Discussion

In this study, we created a novel PRS for ICM-MM, a common form of physical-mental health multimorbidity. Our work demonstrates that combining multiple existing PRSs yields stronger associations with lifetime ICM-MM risk than single trait-specific PRSs. We also demonstrate that including PRS from traits that are not part of our multimorbidity definition further increases the strength of these associations. Multimorbidity is a global public health concern but work on early-life manifestations of multimorbidity that could support identification and prevention are lacking. The novel PRS we constructed could be used in youth cohorts to understand how genetic risk of ICM-MM manifests itself in early life: improving our understanding of the aetiology of this multimorbidity cluster, potentially identifying precursor signals, and highlighting early intervention targets.

The literature on multimorbidity is expanding rapidly, but there is a high degree of heterogeneity in phenotypes under study. This is due, in part, to the canonical definition of ‘two or more long-term conditions’, and study specific decisions about which long-term conditions to include.(1,2) Many pairs of common chronic conditions demonstrate evidence of shared genetic risk,(61) but our ICM-MM phenotype and findings are most similar to those of Baltramonaityte et al (62), who developed a PRS for a multimorbidity phenotype comprising coronary artery disease, T2D and depression. This PRS had a higher OR (1.91, 95%CI = 1.74-2.10) than ICM-MM-PGSs for a similar, but more restricted (n=3 conditions) phenotype, and in a model that was additionally adjusted for age. Zhao et al., (63), also created a PRS for cardiometabolic multimorbidity (two or more of T2D, coronary heart disease and stroke) in UKB with an AUC of 0.62. This was higher than the AUC we found for the ICM-MM-PRSs on ICM-MM, but for a restricted phenotype (n=3 conditions) with no internalising conditions and included age as a component of the model. Other studies that have developed PRS for multimorbidity phenotypes, did not report any comparable metrics (in the case of Gezsi et al (64))), or furthermore explicitly excluded some of the common cardiometabolic conditions we included in our ICM-MM definition (in the case of Zhang et al (65)

Taken together with our findings, the available evidence suggests there is some utility to the use of polygenic risk scores as tools to explore how multimorbidity manifests itself throughout the lifecourse: across a range statistical metrics and similar definitions of multimorbidity, associations between PRSs and multimorbidity phenotypes are modest but persistent. The magnitude of these associations demonstrates a ceiling for associations between polygenic risk scores and ICM-MM or similar multimorbidity phenotypes, regardless of the methods used to derive them.

We highlight that caution is required around communication of the utility of polygenic scores: PRSs alone are not suitable for individual risk prediction or population risk stratification (66), and much of the risk of disease is determined by factors not captured by the genome (67). Recent reviews (68,69) have highlighted the importance of ‘place’ based factors including area-level deprivation and air quality as risk factors for a range of multimorbidity phenotypes, and there is emerging evidence of the role of early-life environmental factors including parental and family environment, and education in the development of midlife hypertension-obesity multimorbidity.(70) Further work is required to elucidate the potential for gene x environmental interactions to influence downstream multimorbidity risk.

## Limitations

We used data from UK BioBank because it is a population-based dataset with known higher prevalence of ICM-MM conditions and rich phenotyping data. However UKB is subject to volunteer bias, and is not representative of the UK population, especially with respect to age, socioeconomic status, or ancestry(71,72) - all factors associated with multimorbidity risk. UKB also contributed to many recent meta-GWAS that would have been ideal sources of summary statistics to construct PRS_TRAIT_. Some but not all studies made available ‘leave out’ summary statistics, excluding UKB, meaning most of the input PRS_TRAIT_ are based on older studies with fewer participants (i.e. weaker instruments). To test the potential of stronger input PRSs, we also carried out a supplemental analysis using 51 PRS created by Genomics PLC: PRS_GPLC_. We found that ICM-MM-PRS_GPLC_ had a stronger association with ICM-MM (OR=1.32, 95%CI:1.28-1.37, AUC = 0.59, 95%CI 0.58-0.60) than single PRS_GPLC_ for ischaemic stroke (the strongest predictor in this validation subset, AUC 0.57, 95%CI .56-.58) and ICM-MM-PRS_TRAIT_ (*p* for difference p=8.13x10^-09^). One of the key limitations of PRS_GPLC_ in our analysis is the lack of PRSs for internalising traits, depression in particular - one of the PRSs most strongly associated with ICM-MM in our PRS_TRAIT_ analysis. We kept PRS_TRAIT_ and PRS_GPLC_ analyses separate because PRS_GPLC_ are only available ‘pre calculated’ in selected datasets and the exact methods required to recreate them are the intellectual property of Genomics PLC.

Somatoform disorders were included in our ICM-MM definition due to high genetic (73), diagnostic(74,75), and symptomatic overlap with depression and anxiety. Our searches did not identify suitable GWAS for the somatoform disorders phenotype that is part of the ICM-MM definition. However, given the strong genetic correlations between somatic disorders and depression and anxiety in particular, (73) it is likely that any potential somatoform PRS_TRAIT_ would likely not be retained in the elastic net regression, as was the case for PRS_TRAIT_ for anxiety.

We also restricted our cohort to individuals with available linked primary care data, reducing the effective sample size, but enhancing the quality of phenotype definition for these conditions, which are commonly diagnosed and managed in primary care.

The Elastic Net regression implemented in PRSmix results in a weighted linear combination of PRS. Since multimorbidity requires a combination of two or more traits to be present, it is possible that modelling non-linear combinations of PRS might improve prediction of MM. Future work could explore the use of machine learning methods to produce non-linear combinations of input PRSs, as well as exploring specific pairwise combinations of ICM-MM conditions.

## Conclusion

Combining multiple polygenic risk scores improves the proportion of variance in liability to ICM-MM explained by genetic variation over single trait specific scores. We have generated a novel polygenic risk score for ICM-MM that can be used to explore manifestations of ICM-MM in individuals at higher genetic risk in external and early-life cohorts. Future work should investigate early-life manifestations of high genetic risk of ICM-MM, as well as environmental risk factors and gene x environment interactions to improve early life recognition and potential interventions for ICM-MM.

## Data Availability

All data produced are available online at

https://biobank.ctsu.ox.ac.uk/crystal/exinfo.cgi?src=accessing_data_guide

## Acronyms

PRS: Polygenic risk score
T2D: Type 2 diabetes mellitus
CKD: Chronic kidney disease
MM: Multimorbidity
ICM: Internalising and CardioMetabolic
HES: Hospital episode statistics
EHR: Electronic health records
AUC: Area under the curve
ROC: Received operator characteristic
LDL-C: Low density lipoprotein C
BMI: Body mass index
BP: Blood pressure

