## Supplemental figures and tables for "Combining polygenic risk scores to understand genetic liability to physical-mental health multimorbidity in UK BioBank"

### SUPPLEMENTAL INFORMATION

**SUPPLEMENTAL INFORMATION	1**

**Supplemental table 1: Characteristics of GWAS used to create PRSTRAIT	2**

**Supplemental figure 1: Associations between PRSTRAIT and internalising and cardiometabolic traits, and ICM-MM	5**

**Supplemental figure 2: Elastic net retained PRSTRAIT weights and performance metrics for ICM-MM in the validation sample	5**

**Supplemental figure 3: Correlations between PRSTRAIT and PRSGPLC	6**

**Supplemental figure 4: Odds ratios for all PRS and ICM-MM in n=44,600 UKB participants	7**

**Supplemental figure 5: AUC for all PRS and ICM-MM in n=44,600 UKB participants	8**

**Supplemental figure 6: Weights for PRSGPLC and prediction metrics in the 50% validation subset	9**

**Supplemental figure 7: Weights for PRSTRAIT and prediction metrics in the GPLC subset	10**

**GWAS references	10**

### Supplemental table 1: Characteristics of GWAS used to create PRS_TRAIT_

| **Reference + data URL / doi** | **Study** | **Trait** | **Sample size** | **Ancestries** | **N cases** | **Variance explained in out of sample prediction** |
| --- | --- | --- | --- | --- | --- | --- |
| (Wray et al. 2018)  10.6084/m9.figshare.21655784 | Psychiatric Genomics Consortium (PGC) - Meta GWAS of seven cohorts. Summary statistics used highlighted in bold)  **PGC29, deCODE, GenScotland, GERA, iPSYCH**,  UK Biobank,  23andMe (discovery sample) | Depression | 143,265 | European | 45,591 | 1.9% (variance explained on liability scale, for full meta analysis) |
| (Meier et al. 2019) | Lundbeck Foundation Initiative for Integrative Psychiatric Research (iPSYCH) study | Anxiety | 31,880 | European | 12,655 | 28% (0.28, SE 0.027) |
| No citation found | - | Somatoform disorder | - | - | - | - |
| (Mahajan et al. 2018)  <https://diagram-consortium.org/downloads.html>  T2D GWAS meta-analysis - Summary of T2D associations, unadjusted for BMI and without UK Biobank subjects | DIAbetes Genetics Replication And Meta-analysis consortium (DIAGRAM): Meta analysis of 32 GWAS (provided sumstats excluding UKB) **BioME**  **deCODE**  **DGDG**  **DGI**  **EGCUT_ExomeCore**  **EGCUT_Human370CNV**  **EGCUT_OmniExpress**  **FHS**  **FUSION**  **GCKD**  **GENOA**  **GERA**  **GoDARTS**  **GOMAP-TEENAGE**  **HPFS**  **INTERACT_coreexome**  **INTERACT_GWAS**  **KORA**  **MESA**  **METSIM**  **MGI**  **NHS**  **NUGENE**  **PIVUS**  **PROSPER**  **RS1**  **RS2**  **RS3**  UK BioBank  **ULSAM**  **UPCH**  **WTCCC** | Type 2 diabetes | 455,313 | European | 55,005 | ~16.3% |
| (Pattaro et al. 2016)  https://ckdgen.imbi.uni-freiburg.de/datasets/Pattaro_2016 | CKDGen: Meta-GWAS from 43 studies | Chronic kidney disease | 117,165 | European | 12,385 | 3.22% |
| (Locke et al. 2015)  https://giant-consortium.web.broadinstitute.org/images/1/15/SNP_gwas_mc_merge_nogc.tbl.uniq.gz | GIANT consortium: Meta GWAS of 114 studies | BMI (proxy for Obesity) | 322,206 | European | NA (quantitative trait) | 6.6% for SNPs with P<5x10^-3^, 21.6% for all HapMap3 SNPs |
| (Willer et al. 2013)  https://csg.sph.umich.edu/willer/public/lipids2013/ | Global Lipids Genetics Consortium Meta analysis of 45 studies | LDL-cholesterol | 188,577 | European, East Asian, South Asian and African ancestry | NA (quantitative trait) | 2.4% |
| (Keaton et al. 2024)  <https://www.ncbi.nlm.nih.gov/projects/gap/cgi-bin/study.cgi?study_id=phs000585.v2.p1> | Meta analysis of 4 cohorts (Sumstats used in bold)  MVP  BioVU  UKB  **ICBP (Large meta analysis of 70 studies)** | (Study reports Systolic and Diastolic blood pressure (SBP/DBP), and pulse pressure (PP), used here to proxy hypertension | 299,024 | European | NA (quantitative trait) | SBP 6.8% DBP 6.83% PP 4.29% |

### Supplemental figure 1: Associations between PRS_TRAIT_ and internalising and cardiometabolic traits, and ICM-MM

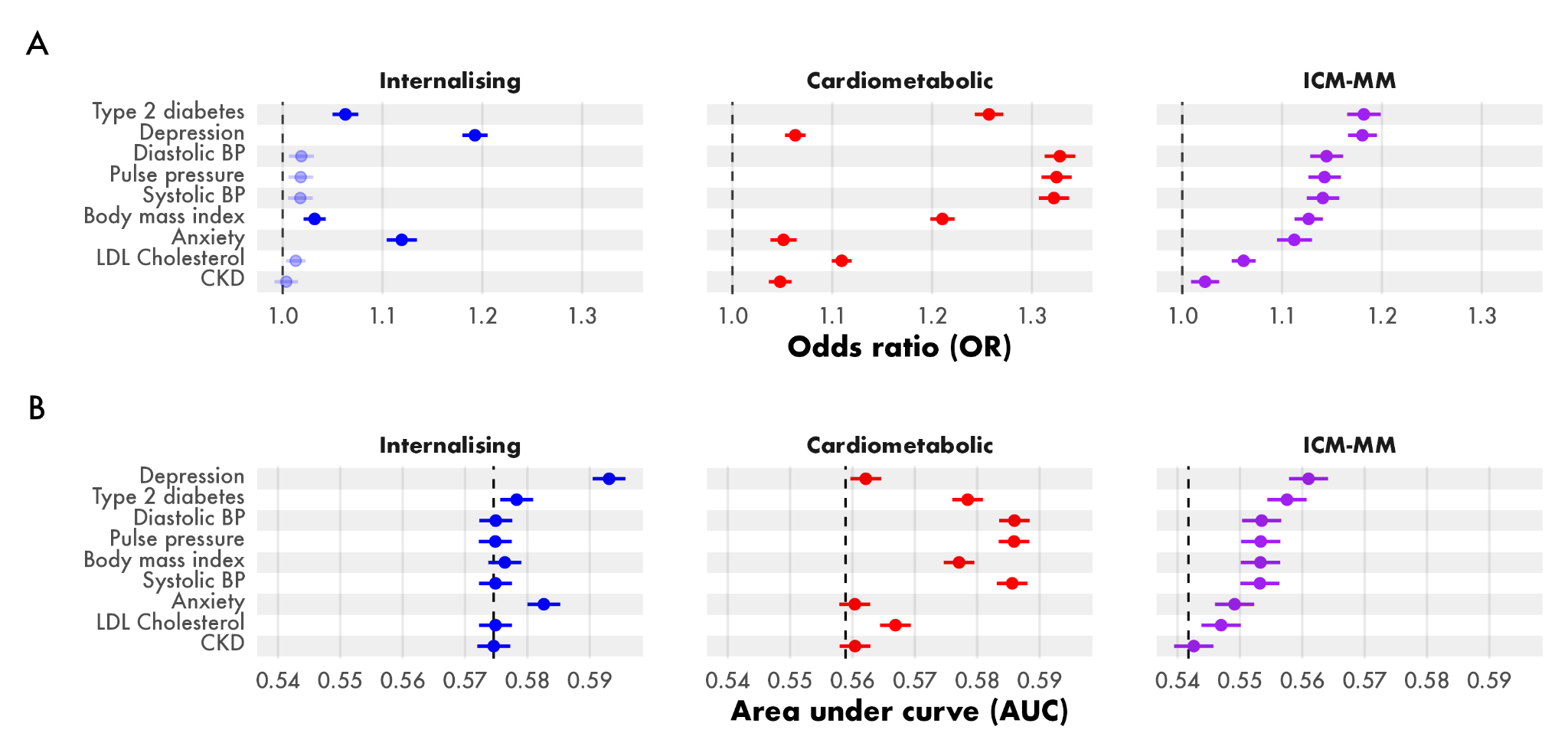

### Supplemental figure 2: Elastic net retained PRS_TRAIT_ weights and performance metrics for ICM-MM in the validation sample

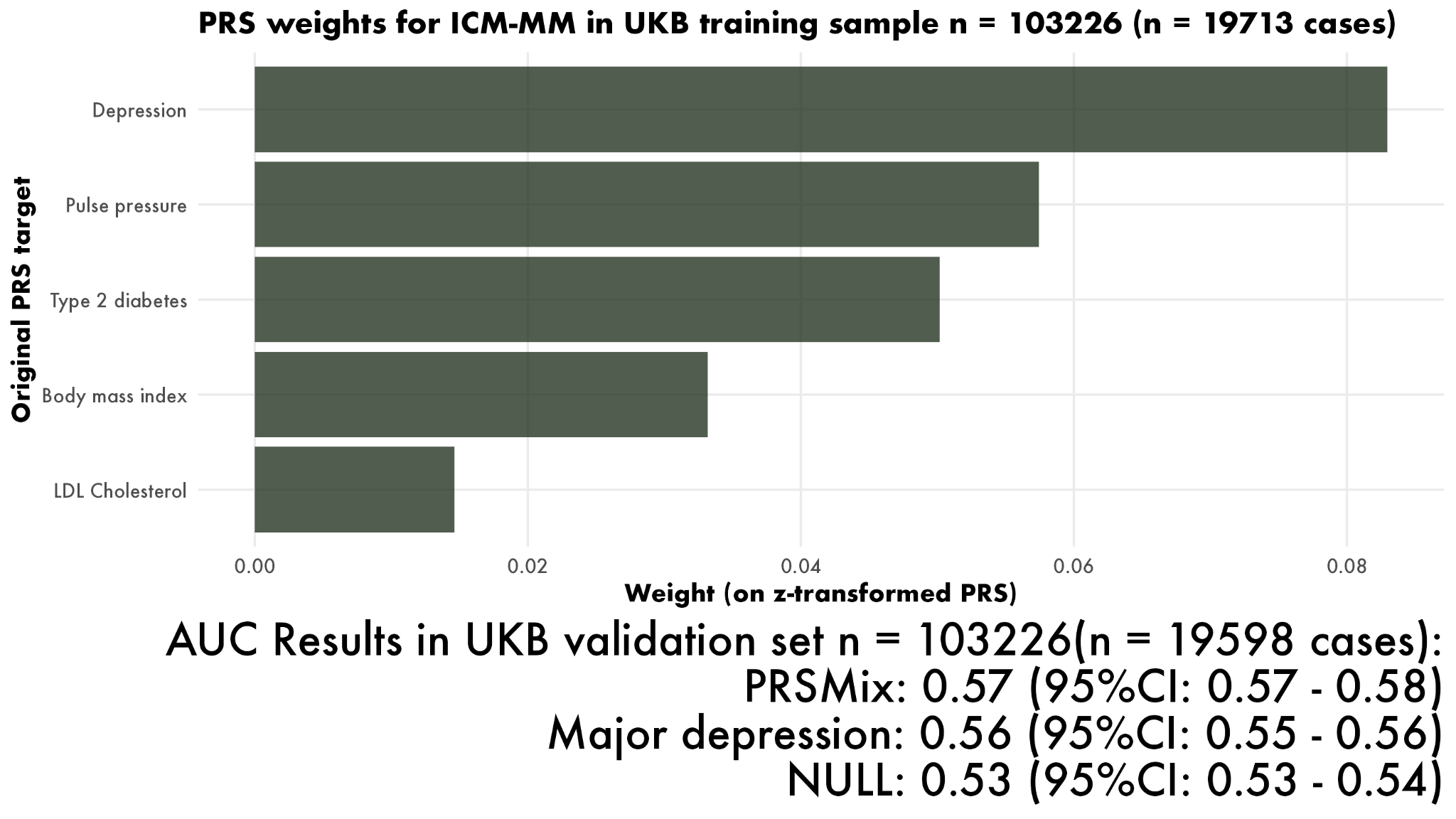

### Supplemental figure 3: Correlations between PRS_TRAIT_ and PRS_GPLC_

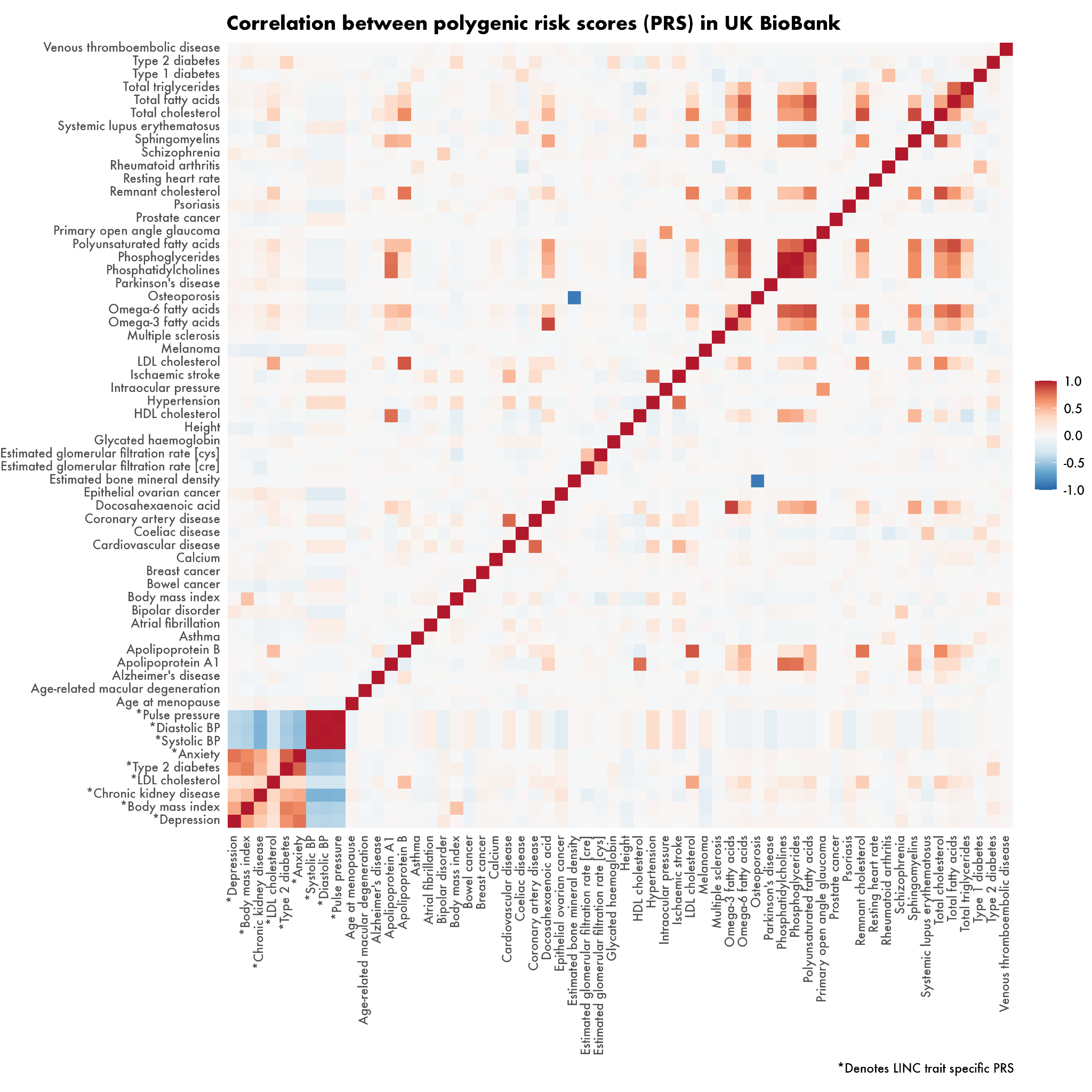

### Supplemental figure 4: Odds ratios for all PRS and ICM-MM in n=44,600 UKB participants

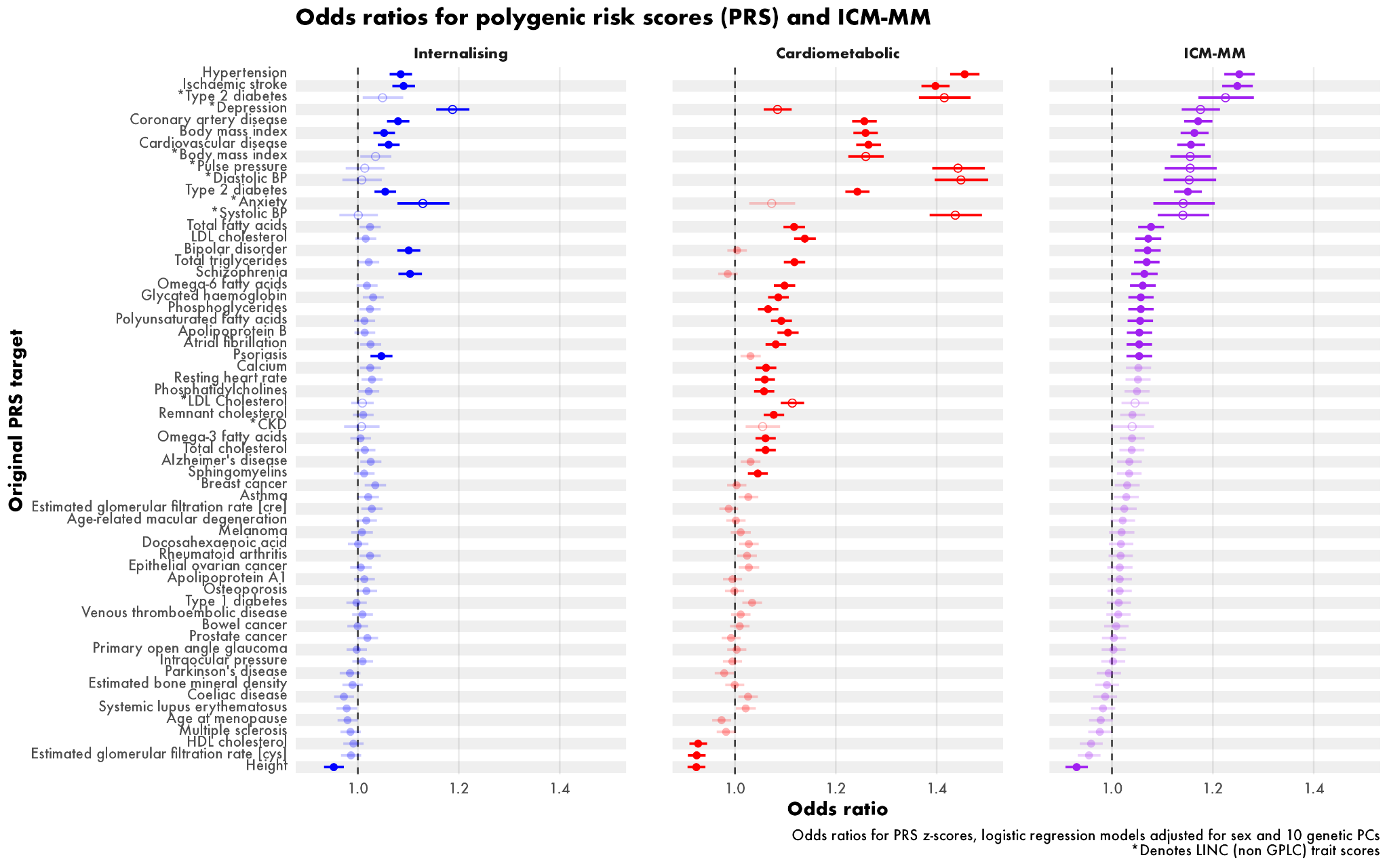

(Figure also includes PRS_TRAIT_ scores for comparison: some participants with PRS_GPLC_ did not meet the inclusion criteria for generating PRS_TRAIT_ hence the number here is lower than for the PRS_GPLC_ analysis n=45,493)

### Supplemental figure 5: AUC for all PRS and ICM-MM in n=44,600 UKB participants

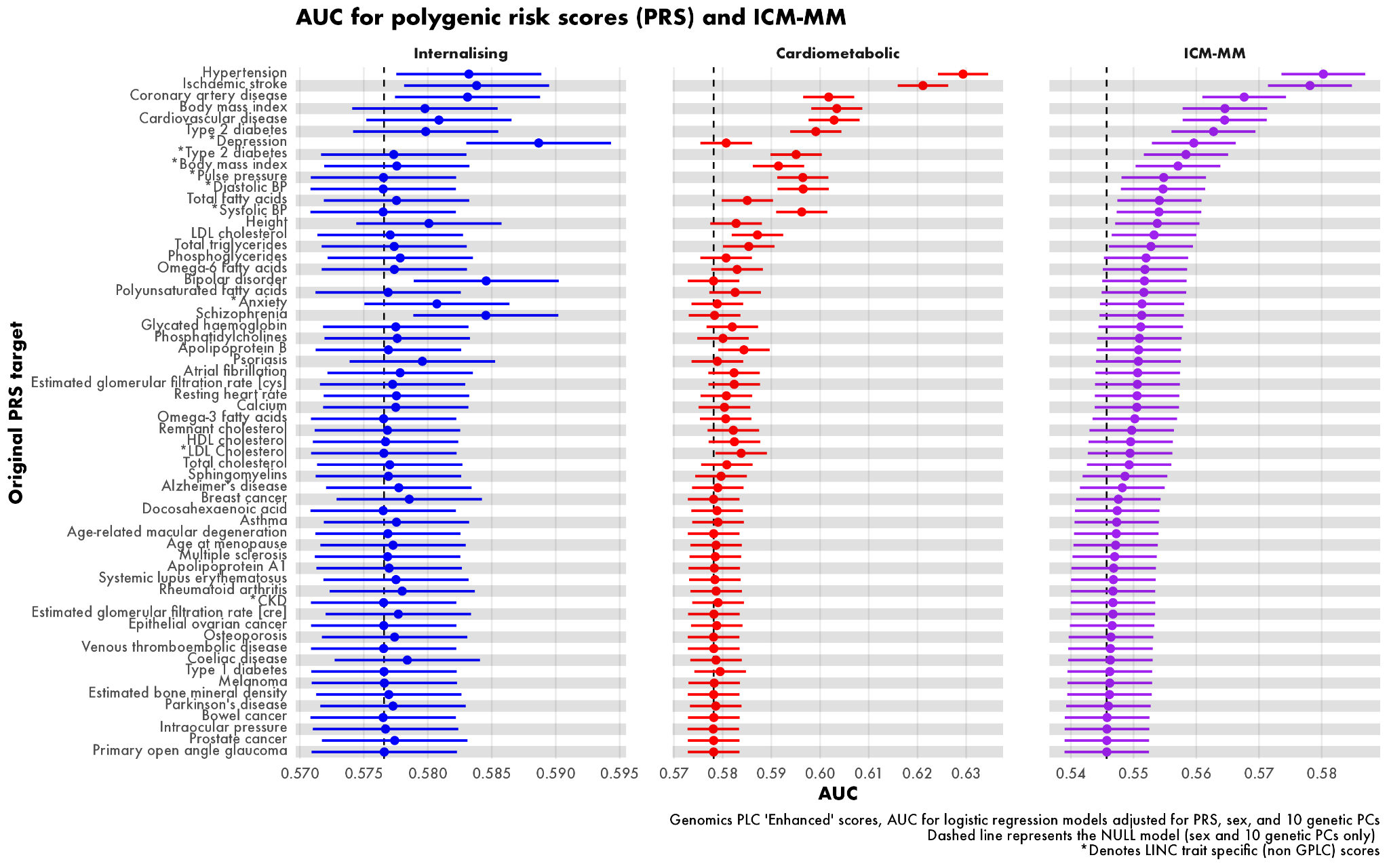

(Figure also includes PRS_TRAIT_ scores for comparison: some participants with PRS_GPLC_ did not meet the inclusion criteria for generating PRS_TRAIT_ hence the number here is lower than for the PRS_GPLC_ analysis n=45,493)

### Supplemental figure 6: Weights for PRS_GPLC_ and prediction metrics in the 50% validation subset

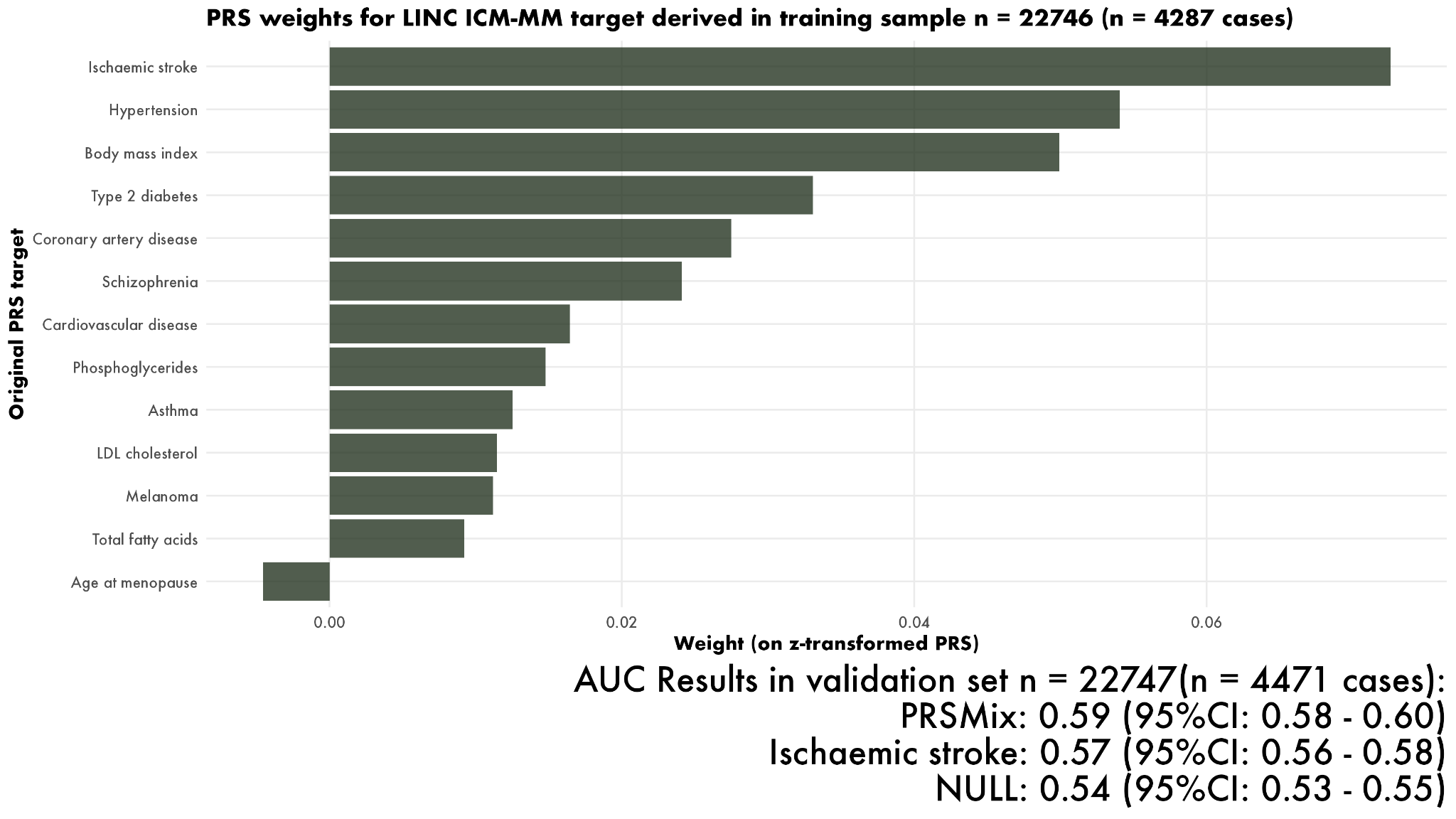

### Supplemental figure 7: Weights for PRS_TRAIT_ and prediction metrics in the GPLC subset

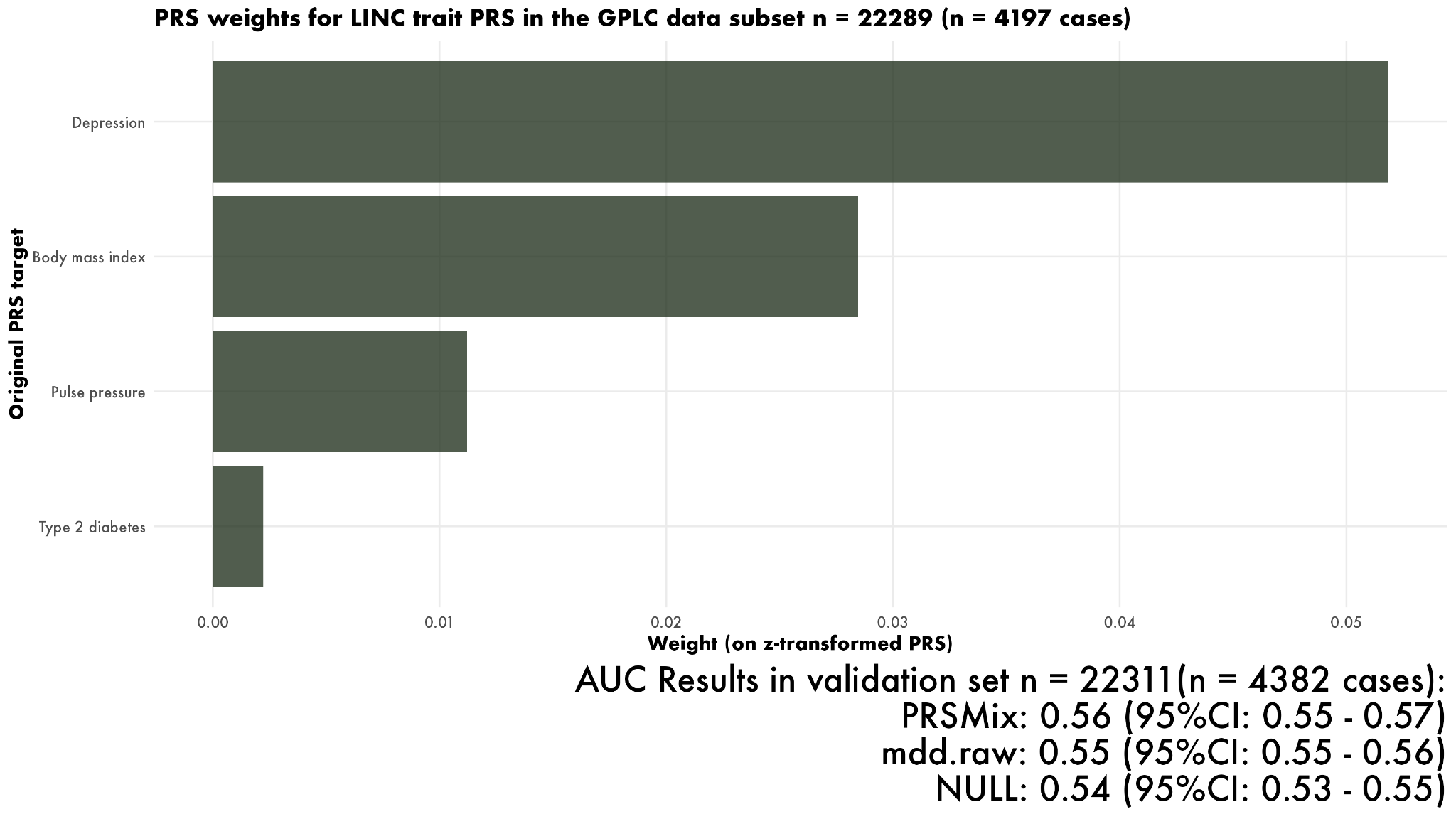

### GWAS references

Keaton, Jacob M., Zoha Kamali, Tian Xie, Ahmad Vaez, Ariel Williams, Slavina B. Goleva, Alireza Ani, et al. 2024. “Genome-Wide Analysis in over 1 Million Individuals of European Ancestry Yields Improved Polygenic Risk Scores for Blood Pressure Traits.” *Nature Genetics* 56 (5): 778–91.

Locke, Adam E., Bratati Kahali, Sonja I. Berndt, Anne E. Justice, Tune H. Pers, Felix R. Day, Corey Powell, et al. 2015. “Genetic Studies of Body Mass Index Yield New Insights for Obesity Biology.” *Nature* 518 (7538): 197–206.

Mahajan, Anubha, Daniel Taliun, Matthias Thurner, Neil R. Robertson, Jason M. Torres, N. William Rayner, Anthony J. Payne, et al. 2018. “Fine-Mapping Type 2 Diabetes Loci to Single-Variant Resolution Using High-Density Imputation and Islet-Specific Epigenome Maps.” *Nature Genetics* 50 (11): 1505–13.

Meier, Sandra M., Kalevi Trontti, Kirstin L. Purves, Thomas Damm Als, Jakob Grove, Mikaela Laine, Marianne Giørtz Pedersen, et al. 2019. “Genetic Variants Associated with Anxiety and Stress-Related Disorders: A Genome-Wide Association Study and Mouse-Model Study: A Genome-Wide Association Study and Mouse-Model Study.” *JAMA Psychiatry (Chicago, Ill.)* 76 (9): 924–32.

Pattaro, Cristian, Alexander Teumer, Mathias Gorski, Audrey Y. Chu, Man Li, Vladan Mijatovic, Maija Garnaas, et al. 2016. “Genetic Associations at 53 Loci Highlight Cell Types and Biological Pathways Relevant for Kidney Function.” *Nature Communications* 7 (1): 10023.

Willer, Cristen J., Ellen M. Schmidt, Sebanti Sengupta, Gina M. Peloso, Stefan Gustafsson, Stavroula Kanoni, Andrea Ganna, et al. 2013. “Discovery and Refinement of Loci Associated with Lipid Levels.” *Nature Genetics* 45 (11): 1274–83.

Wray, Naomi R., Stephan Ripke, Manuel Mattheisen, Maciej Trzaskowski, Enda M. Byrne, Abdel Abdellaoui, Mark J. Adams, et al. 2018. “Genome-Wide Association Analyses Identify 44 Risk Variants and Refine the Genetic Architecture of Major Depression.” *Nature Genetics* 50 (5): 668–81.
